# The Effect of Telenursing on the Quality of Life in Elderly Stroke Survivors: A Triple-Blind Randomized Controlled Trial

**DOI:** 10.1101/2024.12.06.24318582

**Authors:** Elham Nazari, Azin Roumi, Toomaj Sabooteh

## Abstract

**Introduction:** Telenursing is a cost-effective and highly accessible approach that can enhance awareness of care principles, ultimately contributing to the improvement of quality of life (QoL) in stroke patients. This study aimed to assess the impact of telenursing via telephone consultations and video chats on the quality of life of stroke patients.

**Materials and Methods:** This randomized controlled trial included stroke patients discharged from hospitals in Khorramabad city in 2022. A total of 80 patients were selected using non-probability consecutive sampling and randomly allocated to either an intervention group (n=40) or a control group (n=40) using a permuted block randomization method. The study was conducted in a triple-blind design. Research tools included a demographic information form and the Stroke-Specific Quality of Life (SS-QoL) questionnaire developed by Williams et al. (1999). Data were analyzed using independent t-tests and SPSS version 26.

**Results:** The findings indicated a statistically significant difference in the mean QoL scores between the two groups (P<0.001, t = -36.560). The mean difference between the groups was 56.900 ± 1.556. Additionally, the results demonstrated that the telenursing intervention improved various dimensions of stroke patients’ QoL (P < 0.001).

**Conclusion:** Telenursing can be effectively utilized to facilitate the care of chronic patients and improve their quality of life by providing practical and specialized information.

## 1. Introduction

Globally, seven out of the top ten leading causes of death in 2019 were attributed to non-communicable diseases, with ischemic heart disease accounting for 16% of total deaths. Since 2000, ischemic heart disease has shown the highest increase in mortality, rising from approximately 2 million deaths to 8.9 million deaths in 2019. These deaths predominantly occur in adulthood and among active individuals, posing significant economic and social burdens on countries (1). In Europe, ischemic heart disease has declined but still caused 862,000 deaths annually (19% of total deaths) among men and 877,000 deaths among women (2). The elderly are most affected, and the global elderly population, including in Iran, is rapidly increasing. Currently, over 5 million Iranians are elderly, constituting approximately 7.26% of the country’s population. It is estimated that within the next 20 years, the elderly population in Iran will exceed 10% of the total population. Physiological and structural changes in the cardiovascular system associated with aging lead to a higher prevalence of ischemic diseases, particularly myocardial infarction, stroke, and other related conditions (3).

In Iran, ischemic diseases are the leading cause of mortality, contributing to prolonged disability in the elderly. These diseases account for 46% of total deaths and 20–23% of the disease burden in the country (4). The increasing elderly population correlates with the high prevalence of ischemic diseases nationwide (5). Key post-stroke interventions include lifestyle modifications such as smoking cessation, blood pressure control, maintaining a healthy weight, a balanced diet, and encouraging physical activity. The limited time during hospitalization necessitates a multidisciplinary approach to secondary prevention (6). Rehabilitation programs play a crucial role in lifestyle modification, disease prognosis control, and optimization. Over the years, these programs have evolved and demonstrated effectiveness in physical, social, and psychological outcomes for participants (7).

As diseases like ischemic conditions significantly impact patients’ quality of life, the risks associated with surgical interventions in the elderly are inevitably higher (8). Quality of life, a subjective and multidimensional concept, is increasingly recognized as a critical outcome in health and social care research (9). According to the World Health Organization, quality of life pertains to individuals’ perception of their position in life concerning their expectations, standards, and concerns (10). The concept has become a focal point in aging studies due to the growing elderly population and increased longevity. Previous studies have indicated that demographic characteristics (11), chronic disease history (12), and sleep quality (13) are key factors influencing the quality of life in older adults. One major objective for stroke patients participating in rehabilitation programs is improving their quality of life. How elderly stroke survivors adapt and their quality of life post-stroke deserve attention (14). The study by Rejeh et al. (2015) highlighted that the quality of life in stroke patients was suboptimal, necessitating nursing interventions to improve it (5). However, few studies have focused on factors affecting the quality of life in elderly stroke survivors, such as fear of COVID-19, loneliness, resilience, age, gender, chronic disease history, sleep pattern changes, and mental health status (15).

It is crucial for nurses to understand the physical symptoms of patients recovering at home in real time while maintaining face-to-face interaction. Early recognition of physical symptoms enables timely intervention, preventing symptom deterioration (16). The COVID-19 pandemic in 2019 further restricted in-person care, emphasizing the need for innovative strategies to enhance remote rehabilitation programs’ effectiveness (17). The media’s focus on mortality rates, particularly among older adults, combined with governmental isolation measures, quarantine, and physical distancing, negatively impacted elderly mental health (18,19), leading to fear, anxiety, and loneliness (20-22). Studies have reported that all elderly groups experienced depression and anxiety during the COVID-19 pandemic (23). Additionally, long-term restrictive measures have increased loneliness among elderly individuals in nursing homes and private residences. These factors adversely affected their physical health, mental well-being, lifestyle, and quality of life (24-29).

Self-care ability in such circumstances becomes essential, as studies have emphasized the importance of enhancing self-care skills in elderly stroke survivors post-discharge (14,30). Patient education and empowerment in self-monitoring are vital tools supported by telehealth interventions. Remote nursing strategies aimed at self-monitoring chronic diseases have proven effective in modifying high-risk behaviors (31). Among factors influencing quality of life and self-care in elderly stroke survivors, telenursing has emerged as a significant variable. Post-discharge, patients often experience issues like pain, fatigue, limb swelling, activity intolerance, sleep disturbances, wound care challenges, ineffective disease adaptation, and anxiety about their future. Over the past two decades, significant attention has been given to teleconsultations to prevent hospital readmissions. Patient follow-up is essential to address and rectify care needs arising after hospital discharge (32).

Healthcare is a critical sector for any country due to its impact on public health and its significant costs for both developing and industrialized nations. For instance, the United States spends approximately 18% of its GDP on healthcare, more than any other developed country (33). Moreover, a growing population and shortages of physicians and nurses exacerbate healthcare challenges. Telenursing, as an operational tool, has been introduced to enhance healthcare delivery and support, offering substantial changes to healthcare organizations (34). Simultaneously, the increasing elderly population underscores the need for healthcare services and products related to the prevalence of complex medical conditions and chronic diseases (35).

Given the unique national and global conditions arising from the COVID-19 pandemic, the importance of remote care due to health protocols, and the lack of studies on the impact of telenursing on the quality of life in elderly stroke survivors, this research examines the influence of telenursing on their quality of life post-stroke.

## 2. Methodology

### 2.1. Participants and Study Design

This study employed a randomized controlled trial (RCT) design with two groups (intervention and control) to evaluate the impact of remote nursing care delivered through social media-based online education (addressing various aspects of self-care such as dietary adherence, physical activity, personal hygiene, dressing, and medication use) on the quality of life of elderly post-stroke patients.

### 2.2. Study Population and Sampling

The study population comprised stroke patients discharged from hospitals in Khorramabad, Iran, in 2022. A non-probability sequential sampling method was used. Patients were randomly assigned to the intervention or control group using permuted block randomization. A section of a random number table was selected, and single-digit numbers were read. Numbers between 0 and 4 led to the sequence A (control) followed by B (intervention), whereas numbers between 5 and 9 resulted in the sequence B followed by A. This random sequence was implemented in the research setting, and patients were allocated to groups A or B based on their discharge sequence. The allocation process was repeated for each subsequent patient using the table until the corresponding group code (A or B) was selected.

The trial was triple-blinded, meaning that the patients, researchers, and data analysts were unaware of the intervention type. An external individual who was not part of the research team handled the group coding. The initial sample size was estimated to be 33 participants per group using SPSS software version 26. To account for a 20% dropout rate, the final sample size was increased to 40 participants per group.

### 2.3. Inclusion and Exclusion Criteria

Inclusion criteria included: Elderly patients (≥60 years) with a history of stroke at discharge and post-discharge; Patients or their caregivers had access to a mobile phone; Ability to answer phone calls and communicate; Presence of a family caregiver. Exclusion criteria included: Substance abuse leading to non-compliance; Visual or auditory impairments; Cognitive impairment as per the Mini-Mental State Examination (MMSE); Disabling comorbidities.

### 2.4. Intervention

Educational content was developed through a review of relevant literature and clinical guidelines and validated by experts for applicability, relevance, comprehensibility, and utility. Training covered stroke disease, complications, management, risk factors (e.g., smoking, hyperlipidemia, hypertension, weight gain, and stress), dietary and medication regimens, walking techniques (frequency, duration, pulse monitoring within permissible limits based on exercise stress tests), and benefits of walking. Sessions were conducted at the hospital by experienced nurses and university faculty. Each session lasted 40 minutes, followed by a 20-minute Q&A session during which patients practiced tasks under researcher supervision. Any patient difficulties were resolved.

At the last session, each participant’s baseline heart rate (from exercise test results) was documented in an educational form and distributed alongside a booklet. Subsequently, three home visits were scheduled at the end of the first, second, and third months. During these visits, educational reinforcement, monitoring of health behaviors, and encouragement to adhere to the program were provided. Patients practiced walking with researchers to address any issues related to proper technique, pulse control, and physical symptoms during walking. A checklist was completed during each home visit. Patients were also given a phone number for troubleshooting or inquiries.

### 2.5. Online Support (Intervention Group)

After discharge, researchers contacted intervention group patients weekly for one or two sessions via prior coordination between 4 PM and 7 PM. Online education and support were provided, with schedules adjusted based on patient or caregiver preferences. Consultations and referrals to physicians or emergency services were arranged if potential issues arose. Video calls were conducted via available applications (e.g., WhatsApp, Telegram, Instagram) if internet access was sufficient; otherwise, phone calls were used. (36-38).

### 2.6. Outcome Assessment

The quality of life was assessed using the Stroke-Specific Quality of Life (SS-QOL) questionnaire by Williams et al. (1999), which consists of 49 items across 12 dimensions: family roles (3 items), energy (3 items), language (5 items), mobility (6 items), mood (5 items), personality (3 items), self-care (5 items), social roles (5 items), thinking (3 items), upper extremity function (5 items), vision (3 items), and work/productivity (3 items). Scores range from 49 to 245, with higher scores indicating better quality of life (39).

### 2.7. Statistical Analysis

Data were analyzed using SPSS software version 26 after questionnaire scoring and data collection.

## 3. Results

**Table 1** presents the frequency distribution of demographic and background variables categorized by the experimental group. The demographic characteristics of the patients revealed that in the control group, the majority of patients were female (55%), whereas in the intervention group, the majority were male (62.5%). The mean age of patients in the control group was 54.60±17.39 years, while in the intervention group, it was 60±14.88 years. The body mass index (BMI) of most patients in both the control group (52.5%) and the intervention group (87.5%) indicated that they were overweight. The blood type of the majority of patients in both the control group (57.5%) and the intervention group (50%) was blood type A. Further details are provided in Table 1.

**Table 1.**
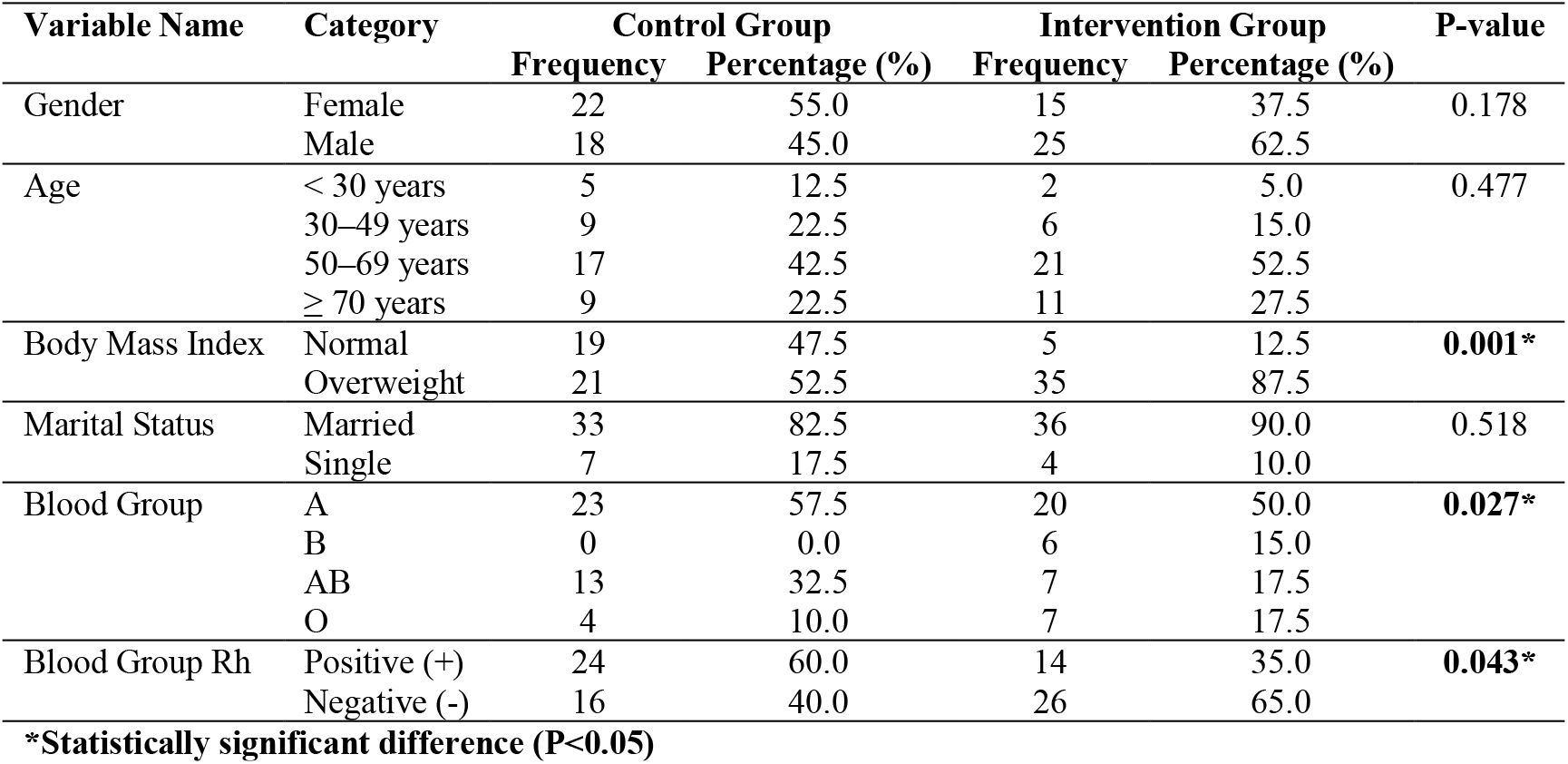
Frequency Distribution of Demographic and Background Variables Among Patients in the Two Study Groups.

In **Table 2**, the descriptive statistics of the research variables are presented separately for the experimental and control groups. The mean score for the **Energy** dimension was 92.59 ± 8.2 in the control group and 77.31 ± 11.1 in the intervention group. The mean score for **Family Role** was 8.45 ± 2.81 in the control group and 12 ± 1.37 in the intervention group. The mean score for **Mood** was 14.05 ± 3.21 in the control group and 19.60 ± 1.70 in the intervention group. The mean score for **Personality** was 8.05 ± 2.15 in the control group and 11.67 ± 1.04 in the intervention group. The mean score for **Social Role** was 13.90 ± 2.72 in the control group and 19.72 ± 1.41 in the intervention group. The mean score for **Mobility** was 17.02 ± 3.41 in the control group and 23.85 ± 1.25 in the intervention group. The mean score for **Thinking** was 8.07 ± 2.57 in the control group and 12.42 ± 1.10 in the intervention group. The mean score for **Upper Limb Function** was 15.15 ± 2.73 in the control group and 19.90 ± 1.17 in the intervention group. The mean score for **Vision** was 9.02 ± 0.50 in the control group and 12.07 ± 1.14 in the intervention group. The mean score for **Productivity** was 8.12 ± 1.63 in the control group and 11.85 ± 1.23 in the intervention group. The mean score for **Language** was 13.90 ± 3.16 in the control group and 20.1 ± 2.65 in the intervention group. The mean score for **Self-Care** was 13.75 ± 2.43 in the control group and 20.1 ± 2.51 in the intervention group. Lastly, the mean score for **Quality of Life** was 138.42 ± 8.56 in the control group and 195.4 ± 32.85 in the intervention group. Further details are presented in Table 2.

**Table 2.** Descriptive Statistics of Research Variables Between the Experimental and Control Groups.

| Variable Name | Group | N | Minimum | Maximum | Mean | SD | Skewness | Kurtosis |
| --- | --- | --- | --- | --- | --- | --- | --- | --- |
| Energy | Control | 40 | 3 | 13 | 8.92 | 2.59 | -0.182 | -0.611 |
|  | Intervention | 40 | 9 | 15 | 11.77 | 1.31 | 0.080 | -0.380 |
| Family Role | Control | 40 | 3 | 15 | 8.45 | 2.81 | 0.251 | -0.302 |
|  | Intervention | 40 | 9 | 14 | 12.00 | 1.37 | -0.558 | -0.138 |
| Mood | Control | 40 | 7 | 20 | 14.05 | 3.21 | -0.077 | -0.298 |
|  | Intervention | 40 | 16 | 23 | 19.70 | 1.60 | -0.187 | 0.072 |
| Personality | Control | 40 | 3 | 13 | 8.05 | 2.15 | -0.164 | -0.078 |
|  | Intervention | 40 | 10 | 14 | 11.67 | 1.04 | 0.002 | 0.671 |
| Social Role | Control | 40 | 10 | 21 | 13.90 | 2.72 | 0.895 | 0.250 |
|  | Intervention | 40 | 17 | 23 | 19.72 | 1.41 | 0.288 | 0.323 |
| Mobility | Control | 40 | 12 | 23 | 17.02 | 3.41 | -0.063 | -1.319 |
|  | Intervention | 40 | 21 | 26 | 23.85 | 1.25 | -0.363 | 0.151 |
| Thinking | Control | 40 | 3 | 15 | 8.07 | 2.57 | 0.671 | 0.425 |
|  | Intervention | 40 | 10 | 14 | 12.42 | 1.10 | -0.099 | 0.886 |
| Upper Limb Function | Control | 40 | 10 | 21 | 15.15 | 2.73 | 0.287 | -0.429 |
|  | Intervention | 40 | 18 | 22 | 19.90 | 1.17 | 0.002 | 0.540 |
| Vision | Control | 40 | 5 | 14 | 9.02 | 2.50 | 0.371 | -1.067 |
|  | Intervention | 40 | 10 | 14 | 12.07 | 1.14 | -0.044 | 0.457 |
| Productivity | Control | 40 | 5 | 11 | 8.12 | 1.63 | -0.322 | -0.484 |
|  | Intervention | 40 | 10 | 15 | 11.85 | 1.23 | 0.734 | 0.798 |
| Language | Control | 40 | 8 | 21 | 13.90 | 3.16 | -0.160 | -0.403 |
|  | Intervention | 40 | 16 | 24 | 20.32 | 1.65 | -0.337 | 0.187 |
| Self-Care | Control | 40 | 8 | 22 | 13.75 | 2.43 | 0.564 | 2.769 |
|  | Intervention | 40 | 17 | 23 | 20.02 | 1.51 | -0.044 | 0.026 |
| Quality of Life | Control | 40 | 121 | 155 | 138.42 | 8.56 | -0.032 | -0.783 |
|  | Intervention | 40 | 182 | 205 | 195.32 | 4.85 | -0.389 | 0.523 |

**Table 3** compares the mean scores of dependent variables between the two experimental groups. The independent t-test revealed statistically significant differences in the mean scores of the following dimensions between the two groups: **Energy dimension:** A significant difference was observed between the groups (P<0.001, t=-6.199), with a mean difference of 2.850 ± 0.459. **Family role dimension:** A significant difference was found (P<0.001, t=-7.156), with a mean difference of 3.355 ± 0.496. **Mood dimension:** A statistically significant difference was noted (P<0.001, t=-9.937), with a mean difference of 5.650 ± 0.568. **Personality dimension:** A significant difference was identified (P<0.001, t=-9.552), with a mean difference of 3.625 ± 0.379. **Social role dimension:** A significant difference was recorded (P<0.001, t=-12.000), with a mean difference of 5.000 ± 0.485. **Mobility dimension:** A statistically significant difference was observed (P<0.001, t=-11.866), with a mean difference of 6.825 ± 0.475. **Thinking dimension:** The analysis showed a significant difference (P<0.001, t=-9.813), with a mean difference of 4.350 ± 0.443. **Upper limb functioning dimension:** A significant difference was found (P<0.001, t=-10.105), with a mean difference of 4.750 ± 0.470. **Vision dimension:** The difference between the groups was statistically significant (P<0.001, t=-7.005), with a mean difference of 3.050 ± 0.435. **Productivity dimension:** A significant difference was identified (P<0.001, t=-11.507), with a mean difference of 3.725 ± 0.323. **Language dimension:** A statistically significant difference was observed (P<0.001, t=-11.390), with a mean difference of 6.425 ± 0.564. **Self-care dimension:** The analysis revealed a significant difference (P<0.001, t=-13.845), with a mean difference of 6.275 ± 0.453. **Quality of life dimension:** A highly significant difference was noted (P<0.001, t=-36.560), with a mean difference of 56.900 ± 1.556. Further details can be found in Table 3.

**Table 3.** Comparison of Dependent Variables Between the Two Experimental Groups.

| Variable Name | Mean Difference | Standard Deviation of Difference | t-Statistic | P-Value | 95% Confidence Interval |
| --- | --- | --- | --- | --- | --- |
| Energy | -2.850 | 0.459 | -6.199 | P < 0.001 | -3.765 to -1.934 |
| Family Role | -3.355 | 0.496 | -7.156 | P < 0.001 | -4.537 to -2.562 |
| Mood | -5.650 | 0.568 | -9.937 | P < 0.001 | -6.781 to -4.518 |
| Personality | -3.625 | 0.379 | -9.552 | P < 0.001 | -4.380 to -2.869 |
| Social Role | -5.825 | 0.485 | -12.000 | P < 0.001 | -6.791 to -4.858 |
| Mobility | -6.825 | 0.475 | -11.866 | P < 0.001 | -7.970 to -5.679 |
| Thinking | -4.350 | 0.443 | -9.813 | P < 0.001 | -5.232 to -3.467 |
| Upper Limb Function | -4.750 | 0.470 | -10.105 | P < 0.001 | -5.685 to -3.814 |
| Vision | -3.050 | 0.435 | -7.005 | P < 0.001 | -3.916 to -2.183 |
| Productivity | -3.725 | 0.323 | -11.507 | P < 0.001 | -4.369 to -3.080 |
| Language | -6.425 | 0.564 | -11.390 | P < 0.001 | -7.548 to -5.302 |
| Self-Care | -6.275 | 0.453 | -13.845 | P < 0.001 | -7.177 to -5.372 |
| Quality of Life | -56.900 | 1.556 | -36.560 | P < 0.001 | -59.998 to -53.801 |

## 4. Discussion

In this study, we aimed to evaluate the impact of a cost-effective approach—remote nursing through telephone consultations and video chats—on improving the quality of life for elderly patients with stroke. Patients with chronic diseases often experience low quality of life. Our findings indicate that providing remote nursing via telephone and video chat consultations can significantly enhance the quality of life for stroke patients.

Stroke is a major health issue in modern societies. The incidence of first-ever stroke in Iran is approximately 139 cases per 100,000 individuals. Due to advancements in the diagnosis and treatment of diseases, the number of stroke survivors is increasing. Upon hospital discharge, about 80% of stroke patients rely on family members and caregivers for daily activities (40). Besides the mortality associated with stroke, its related disabilities impose a significant economic burden on the healthcare system, costing approximately $57 billion annually (41). Remote nursing is recognized as one of the primary services for managing chronic patients at home (42). This technology facilitates quick access to better healthcare services at lower costs, easier access to specialized professional skills, and overall enhancement in the quality of healthcare delivery (43). Given the chronic nature of stroke, supporting family members is critical in increasing their efficiency in caregiving.

In Iran, based on the experiences of stroke patients and their families, most caregivers report facing numerous challenges due to inadequate social and financial support, lack of educational programs, and rehabilitation services, as well as physical and psychological burdens. The lack of adequate support and education to address the disease, financial difficulties, and familial blame associated with the illness are among the challenges faced by patients and their caregivers in our society. However, the active role of nurses in educating patients and their families about managing and caring for chronic illnesses is often referred to as the missing link in comprehensive care responses. Research has shown that involving family members in patient care is an essential part of nursing programs. They can provide the necessary knowledge, skills, and support to maintain the quality of care at home (44).

Follow-up care can be facilitated through in-person clinic visits or scheduled home visits. In the case of stroke, due to the importance of long-term follow-up, the approach must be cost-effective and feasible for a large number of patients. Today, telephone consultations enable nurses to perform activities such as patient monitoring, education, data collection, nursing interventions, pain management, and family support. Unlike in-person visits, remote nursing technology allows for effective care interventions in a shorter time frame. Patient care can shift from hospital-centered to community-centered and from care-centered to patient-centered.

Some studies have shown that empowering stroke caregivers through the provision of necessary educational programs improves the quality of life for patients. Furthermore, training caregivers of stroke patients based on the family-centered empowerment model enhances their knowledge and self-esteem, thereby improving their role and contribution to effective care (45). Unfortunately, stroke patients currently lack the support of social organizations or associations in Iran. Many of these patients require essential services to meet their nutritional needs, manage urinary incontinence, improve speech, prevent pressure sores, and more. However, the burden of care falls on caregivers. Therefore, remote nursing can be proposed as a cost-effective and practical solution to improve the quality of life for stroke patients.

## 5. Conclusion

Considering the circumstances arising from the COVID-19 pandemic in recent years and the importance of healthcare for elderly individuals who have experienced a stroke, the adoption of novel approaches to patient care and the education of patients and their caregivers has gained particular significance. Tele-nursing is one such innovative method that, during a pandemic, can support the healthcare system both economically (cost-effectiveness) and in terms of public health (improving patients’ quality of life). Therefore, it can be stated that telehealth services can facilitate chronic patient care and enhance patients’ quality of life by providing practical and specialized information.

## Data Availability

All data produced in the present study are available upon reasonable request to the authors

## Acknowledgements

We are grateful for the financial support and resources provided by Lorestan University of Medical Sceinces, without which this research would not have been possible. We would like to express our sincere gratitude and appreciation to Dr. Monireh Kamali, Amir Hoseini, Dr. Yunes Ghasemi, and Dr. Nasrin Sharifi for their valuable collaboration. This research was conducted in partnership with Shiraz University of Medical Sciences and Qom University of Medical Sciences.

## Author Contribution

The development of this manuscript was a collaborative effort by all authors. **E.N**. supervised the study design and coordinated the data collection process. **A.R**. carried out the data analysis and prepared the initial draft of the manuscript. **T.S**. critically reviewed, edited, and provided overall supervision. All authors participated in reviewing and approving the final version of the manuscript.

## Ethical Considerations

This study was conducted in accordance with the ethical principles outlined in the Declaration of Helsinki. Ethical approval was obtained from the Ethics Committee of Lorestan University of Medical Sceinces. Written informed consent was obtained from all participants after explaining the study objectives, procedures, and their rights, including the right to withdraw at any stage without any impact on their treatment or care. Confidentiality of participants’ information was strictly maintained, and all data were anonymized for analysis. Additionally, care was taken to ensure that the intervention did not impose any undue burden or harm on the participants.

